# Beyond maternal health: a gendered analysis on the role of publicly-funded health insurance in expanding health service utilisation and financial protection for Indian women

**DOI:** 10.1101/2025.06.13.25329614

**Authors:** Susanne Ziegler, Swati Srivastava, Divya Parmar, Sharmishtha Basu, Nishant Jain, Manuela De Allegri

**Affiliations:** Heidelberg Institute of Global Health, Medical Faculty and University Hospital, Heidelberg University Im Neuenheimer Feld 130.3, 69120 Heidelberg, Germany; Deutsche Gesellschaft für Internationale Zusammenarbeit (GIZ) GmbH Friedrich-Ebert-Allee 32+36, 53113 Bonn, Germany; Department of Population Health Sciences, School of Life Course and Population Sciences, King’s College London, Weston Education Centre, Cutcombe Road, London SE5 9RJ, United Kingdom; Deutsche Gesellschaft für Internationale Zusammenarbeit (GIZ) GmbH B5/1 Safdarjung Enclave, 110029 New Delhi, India

## Abstract

**Introduction:** There is a lack of evidence regarding the role of universal health coverage (UHC) schemes, such as public-funded health insurance (PFHI), on women’s utilization of health services and financial protection, particularly for conditions beyond maternal care. To address this gap, we conducted a gender analysis of health care utilization and out-of-pocket expenditure (OOPE) among poor and vulnerable women in India, focusing on India’s first large national PFHI scheme, *Rashtriya Swasthya Bima Yojana* (RSBY).

**Methods:** We analyzed data from a cross-sectional household survey in eight Indian states to compare the utilization and OOPE of women and men among RSBY-eligible families. We used bivariate analysis, multivariate logistic and GLM regressions, controlled for demographic and socio-economic characteristics and examined maternal health and non-maternal health services separately.

**Results:** Independently of enrollment in RSBY, women had a higher use of hospitalization services than men, but only when services for maternal health were included in the analysis (women: 4%, men: 3.23%, p < 0.001). Enrollment in RSBY did not influence the decision to be hospitalized for either men or women. However, it led to reduced OOPE for women who were hospitalized for services other than maternal health (coef = -0.26; p = 0.084).

**Conclusion:** The findings highlight the potential benefits of PFHIs in providing financial protection for women beyond maternal health. To further improve access to health care for poor and vulnerable women, it is imperative that their health care needs are considered throughout their lifecycle when healthy policy interventions are designed and implemented. We recommend further research on the gendered effects of PFHIs. Findings should feed into the reform processes of PFHIs in India and other low- and middle-income countries, ensuring that these schemes are gender-responsive, universally designed and implemented and ultimately reach all poor and vulnerable individuals.

## 1. Introduction

Universal health coverage (UHC) entails that all people should have access to the essential health services they need without suffering from financial hardship (1). Many people in low- and middle-income countries (LMICs) face difficulties in accessing such services due to persisting inequities in wealth, ethnicity, education, geographic location and sociocultural factors (2–4). This is especially true for women as gender norms and power relations that persist in many societies influence women’s ability to access health care services and ultimately their health outcomes (5). To improve access to health services and increase financial protection in low-income settings, many LMICs have introduced publicly-funded health insurance (PFHI) schemes. These schemes are usually introduced as universal schemes, meaning that they target the entire population, including the poor and most vulnerable, who are often excluded from other forms of insurance and social health protection coverage. Examples of such schemes include statutory health insurance in Indonesia and Thailand (6–8).

PFHI has been recommended as one of the most equitable routes to achieving UHC (9,10). Systematic reviews highlight that PFHIs have a positive effect on improving access to health services and reducing out-of-pocket expenditure (OOPE) for health care (11,12). Nonetheless, specific evidence on the benefits accrued by women, both in terms of increased health service utilisation and financial protection, remains extremely limited. This lack of evidence may be derived from the assumption that a well-functioning health system progressing toward UHC will inherently be equitable and gender-neutral (13,14). The limited evidence available regarding the potential effects of PFHIs on women focuses mainly on their utilisation of maternal and child health (MCH) services and confirms that different types of health insurance improve women’s access to these services (15–18). This focus is not surprising, as health policy reforms, health care delivery systems and research have long prioritised women’s reproductive capacities with significant improvements in MCH outcomes: for instance, the global maternal mortality ratio dropped by 38% between 2000 and 2017 (19).

Nonetheless, women’s health needs are much broader than just MCH and reproductive health, and this should be acknowledged throughout women’s entire lifecycle. This is especially important because, for example, non-communicable diseases (NCDs) have become the leading cause of death among women, accounting for 73% of all deaths of women worldwide (3). In addition, women living in more gender-inequitable societies are more likely to suffer from the double burden of infectious diseases (IDs) and NCDs as their dependency on men and other household members often hinders them from seeking treatment (20).

The available evidence examining the benefits accrued by women enrolled in different types of health insurance beyond MCH services is limited to a handful of studies. Using data from the 2002-04 World Health Survey in 48 LMICs, El-Sayed et al. (2015) revealed that health insurance improved the likelihood of women receiving treatment for NCDs, but the study did not differentiate between different types of insurance, such as publicly-funded, private or community-based health insurance (21). Another study by Samarakoon and Parinduri (2015) found that enrolment in a PFHI in Indonesia did not increase women’s use of preventive and curative care (18). To the best of our knowledge, no study to date has examined a reduction in OOPE specifically for women enrolled in PFHI accessing services beyond MCH. This implies that we do not have a full understanding of the potential for financial protection that PFHI can have for this population group.

The gaps observed at the global level are also reflected in the Indian context. India has a longstanding tradition of not only implementing PFHIs at central and state levels, but also of initiating health programs specifically targeted at women with a focus on MCH services, which resulted, among others, in the reduction of the maternal mortality ratio by 59% between 2000 and 2017 (19). There is less focus on other services, although, for example, the prevalence for NCDs and IDs is higher for Indian women (63 per thousand persons and 8.32 respectively) than for men (47 and 8.2 respectively) (22,23).

PFHIs in India provide a broad range of services, including for MCH, NCD and ID, and mainly cover the costs of hospitalisations. The *Rashtriya Swasthya Bima Yojana* (RSBY), operational from 2008 to 2018, is a PFHI which was introduced in order to improve access to health care services and reduce OOPE for poor and informally employed population groups. Available research on RSBY provides mixed results regarding women: overall enrolment in RSBY was largely gender-neutral, and households headed by women were more likely to enrol (24–26). Numerous studies conclude that the scheme has had limited or no success in providing financial protection to beneficiaries and has even increased OOPE, but the results do not differentiate between men and women (27–32). Research also suggests that access to health services improved for women under RSBY: once enrolled, women used services more often than men (27,33), yet studies found that women used RSBY mainly for MCH services like deliveries (34,35). This led to reduced infant and under-five mortality especially among urban poor households (35). Other studies have specifically analysed the utilisation of MCH services under RSBY and revealed that RSBY reduced OOPE related to MCH services, but revealed also that effects could have been higher if beneficiaries had been better informed about the entitlements and benefits of the scheme (34,36,37). To our knowledge, literature on the effect that PFHI can have on women when they utilise non-MCH services is lacking for India, including for RSBY.

Our brief review of the literature points to the need to build a more robust body of evidence on the benefits accrued by women given their enrolment in a PFHI, specifically measured in relation to an increased utilisation of health care services and increased financial protection. This is essential to inform more gender-sensitive health policies and remove financial barriers that women face when accessing services, especially in light of India’s shift in disease burden and the rising prevalence of NCDs among women (38).

Our work is thus set against this background and aims to fill this gap in knowledge by performing a gendered analysis of health care utilisation and related OOPE under RSBY across eight Indian states. We achieved this by examining the extent to which RSBY resulted in the same benefits for men and women, measured in terms of increased health service utilisation and reduced OOPE. In line with our intention to move beyond existing analyses of PFHI, which focus mostly on MCH matters, we examined the effect of the scheme on service use, both including and excluding the utilisation of maternal health services. RSBY was subsumed under the larger *Pradhan Mantri Jan Arogya Yojana* (PM-JAY) in 2018, but it still presents an interesting research opportunity as there are many similarities in the implementation modalities of RSBY, PM-JAY, as well as other PFHIs active in other states. We trust that useful lessons for policy makers and implementers can be learned, given RSBY’s originality and the wide geographical scope of our sample. In the following, we employed the term *sex* to refer to distinctions that arise from the biological differentiation of being male or female, while *gender* was used to describe societal roles.

## 2. Materials and methods

### Study setting

The benefit package of RSBY covered approximately 1500 specific procedures and hospitalisation and day care services including pre-existing conditions (39,40), but excluded outpatient, preventive and primary care. MCH services such as institutional deliveries, antenatal check-ups, and diagnostics and treatments after birth were part of the benefits package (40). Households were able to enrol in RSBY if they were living below the poverty line, were registered for the public works programme *Mahatma Gandhi National Rural Employment Guarantee Act*, the subsidised food programme *Antyodaya Anna Yojana* or had an RSBY card in the past (41). A maximum of five members per household were able to enrol including the head of household, the spouse and up to three dependents. To access benefits, eligible households had to enrol for RSBY, i.e., they had to pay an annual registration fee of 30 INR (approx. 0.45 USD in 2016), visit an enrolment station and provide personal information such as names, fingerprints and photos. They were then issued a chip-based biometric insurance card on the spot, by which they could access RSBY benefits from public and empanelled private hospitals across India (33,42).

### Data sources and sampling

The study uses secondary data from a cross-sectional household survey conducted in 2014 as part of a larger evaluation of RSBY in the Indian states of Bihar, Gujarat, Kerala, Mizoram, Tripura, Uttarakhand, Uttar Pradesh and West Bengal. The eight study states were purposively selected in consultation with the Indian central and state governments to encompass the wide range of demographic and socio-economic characteristics of Indian states, as well as to reflect the varying levels of RSBY implementation progress across states. The detailed sampling strategy is described in Ziegler et al (26). The individuals who participated in this study were selected through a combination of multi-stage purposive and random sampling methods. In Gujarat, Kerala, Mizoram and Tripura, around 1000 households each were interviewed, and in Bihar, Uttarakhand, Uttar Pradesh and West Bengal, around 900 each. This resulted in a total study sample consisting of 7 609 households with 36 665 individuals. The secondary data were accessed by the authors for research purposes on 1st October 2019. The authors had no access to information that could have identified individual participants during or after data collection. Figure 1 depicts the pathway to identify the composition of the final sample.

**Fig. 1:**
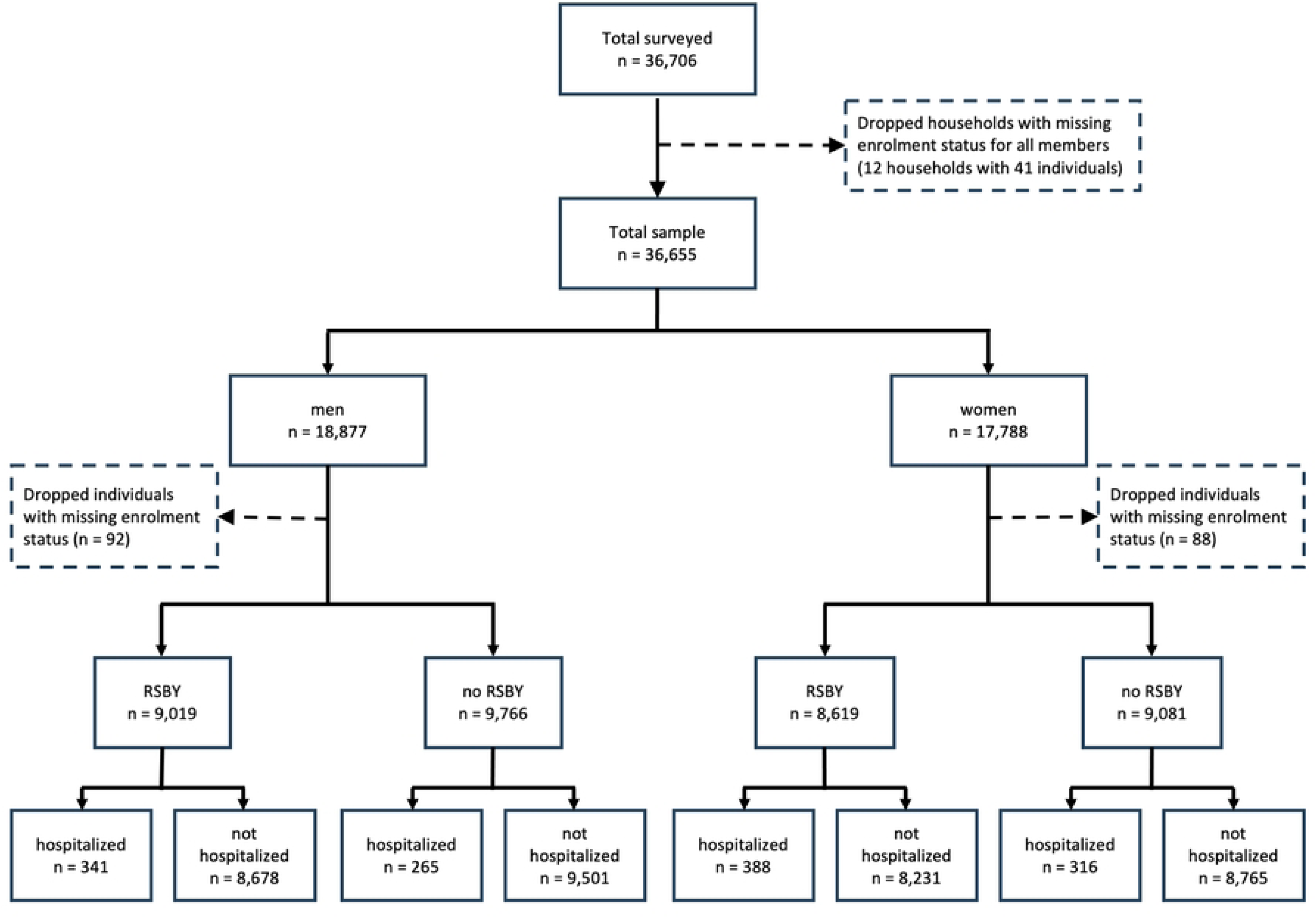
Composition of the study sample. Source: own depiction.

### Variables and their measurement

Table 1 provides an overview of the variables used in the analyses with their unit of measurement and distribution in the sample. The analysis took into consideration two outcome variables: (1) a binary variable *hospitalisation* denoting whether an individual was hospitalised (*yes*) or not (*no*) in the last 365 days; (2) *OOPE*, a continuous variable that denoted the OOPE (in INR) associated with the respective hospitalisation in the last 365 days. OOPE included direct medical expenditures, namely consultation fees, diagnostic tests, and medicines, and non-medical expenditures such as food and transportation costs. We found a strong right-tailed distribution and extreme outliers. As this is problematic for the regression models, we symmetrically winsorised the OOPE variable by replacing the lower and upper 3% of outliers with the respective lowest and highest values of the subsample (at the 3^rd^ and 97^th^ percentiles, respectively) (43). This approach allowed us to keep all observations and prevent outliers from distorting the findings of the regression models. It has previously been applied for analyses with similar challenges (6,44). The variables *sex* (male or female) and *enrolment in RSBY* (yes or no) were the key exposure variables. To account for potential confounders, we included a set of covariates that the captured socio-economic, demographic and geographic characteristics of individuals and households.

**Table 1:**
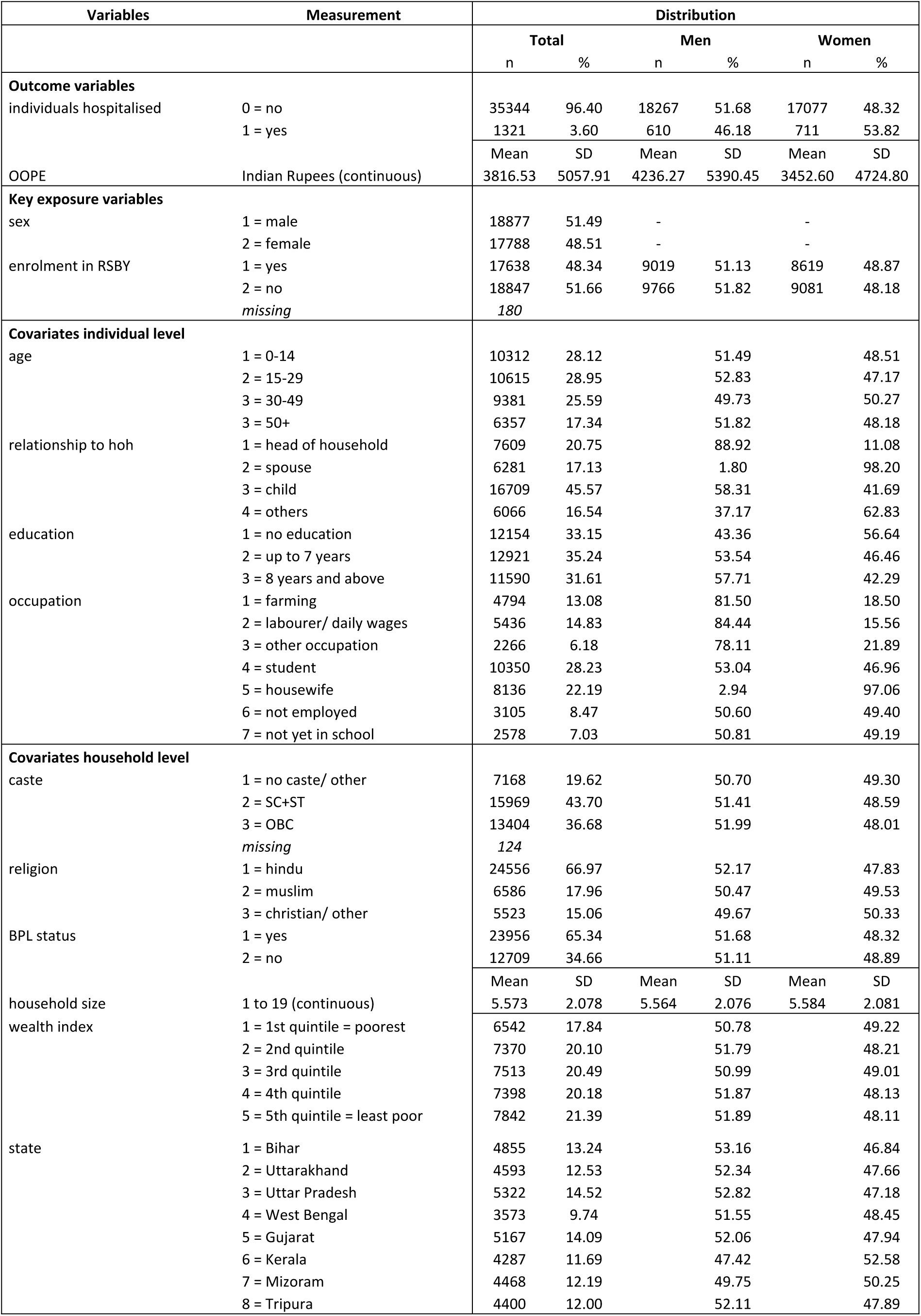
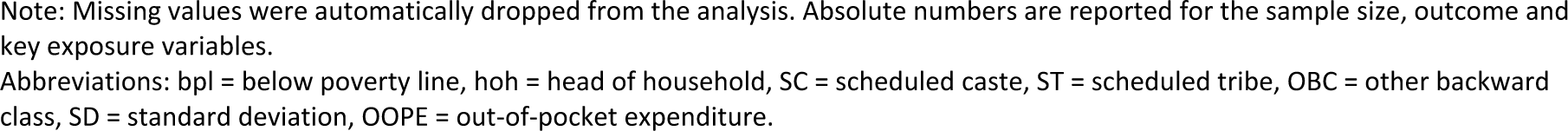
Used variables, their measurement and their distribution in the sample (in total and disaggregated by sex)

Table 2 includes an overview of the sex-specific distribution of hospitalisation services sought after by persons who participated in this study. For our analysis, we summarised deliveries (n=93) and Caesarean-sections (n=10) as maternal health (MH) services (n=103).

**Table 2:** Sex-specific distribution of hospitalisation services.

|  |  | Maternal health Services |  | Other services |  |  |  |  |  |  |  |  |  |  |  |  |  |  |  |  |  |  |  | Total |
| --- | --- | --- | --- | --- | --- | --- | --- | --- | --- | --- | --- | --- | --- | --- | --- | --- | --- | --- | --- | --- | --- | --- | --- | --- |
|  |  | C-section | Delivery | Accident/ injuries | Arthritis | Asthma | Cancer | Cardiac treatment | Cataract | Cholera | Dengue | Diabetes | Diarrhea | Fever | Gastric treatment | Gynecological treatment | Hypertension | Kidney | Malaria | Paralysis | Pneumonia | Typhoid | Others* | Total |
| Men | n | - | - | 75 | 15 | 23 | 15 | 46 | 16 | 11 | 3 | 10 | 32 | 30 | 10 | - | 15 | 20 | 29 | 13 | 5 | 27 | 215 | 610 |
|  | % | - | - | 12.3 | 2.46 | 3.77 | 2.46 | 7.54 | 2.62 | 1.8 | 0.49 | 1.64 | 5.25 | 4.92 | 1.64 | - | 2.46 | 3.28 | 4.75 | 2.13 | 0.82 | 4.43 | 35.25 | 100 |
| Women | n | 10 | 93 | 32 | 17 | 17 | 52 | 22 | 17 | 7 | 11 | 14 | 27 | 31 | 10 | 50 | 19 | 12 | 20 | 5 | 7 | 34 | 204 | 711 |
|  | % | 1.41 | 13.08 | 4.5 | 2.39 | 2.39 | 7.31 | 3.09 | 2.39 | 0.98 | 1.55 | 1.97 | 3.8 | 4.36 | 1.41 | 7.03 | 2.67 | 1.69 | 2.81 | 0.7 | 0.98 | 4.78 | 28.69 | 100 |
| Total | n | 10 | 93 | 107 | 32 | 40 | 67 | 68 | 33 | 18 | 14 | 24 | 59 | 61 | 20 | 50 | 34 | 32 | 49 | 18 | 12 | 61 | 419 | 1321 |

\* The “Others” category includes services which either have a very low sample size or were categorised as “others” already during the data collection process.

### Analytical approach

To estimate the association between RSBY enrolment, health service utilisation and OOPE, we differentiated between hospitalisations for all services, including MH services, and hospitalisations for non-MH services. This means that we ran each analysis twice: first, on the sample that included hospitalisations for all services, and second for the sample including only non-MH services.

We used descriptive statistics to illustrate the distribution of the outcome variables and to identify how many individuals (both men and women) were hospitalised. We conducted bivariate analyses to explore the distribution of the covariates among hospitalised and non-hospitalised individuals. For the OOPE analyses, we only included individuals who had a hospitalisation in the last year (i.e., 1321 individuals). We did not consider individuals who were hospitalised twice (n=21) or thrice (n=1) in the last year due to too many missing values. We calculated the mean and standard deviation (SD), and applied t-tests and analyses of variance (ANOVA) to examine differences among the variable categories.

We estimated two regression models: first, a multivariate logistic regression for *hospitalisation,* where we measured the likelihood of an individual, situated within a given household, being hospitalised during the past year. The model was expressed as follows:

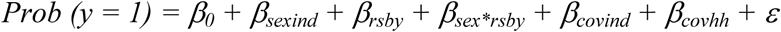

*y* was the binary outcome variable taking the value ‘1’ for the hospitalisation of an individual and ‘0’ for no hospitalisation in the past year. *sexind* (sex of the individual) and *rsby* (enrolment in RSBY) were the main exposure variables as we assumed that the hospitalisation depended on the individual’s sex and enrolment status in RSBY. We also included the interaction term *sex*rsby* to understand if, and to what extent, the association between RSBY and hospitalisation was different for men and women. We included a wide set of socio-economic characteristics at the individual (*covind*) and household (*covhh*) levels to control for possible confounders.

Second, as the outcome OOPE was highly skewed, we fitted a generalised linear model (GLM) gamma regression with a log link function. GLMs with gamma regression modelling techniques provide more accurate estimates of population means of health care expenditure than other models and they accommodate skewness (45,46). The model was expressed as follows:

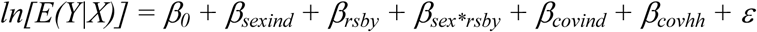

ln[E(Y|X)] was the log link function for the outcome OOPE. We included the same exposure variables, covariates and interaction term as in the first model. For all analyses, standard errors were clustered at the household level as individuals living in the same household share similar characteristics. The Wald helped us to test variable inclusion and the Likelihood Ratio Test (LRT) to test for interactions and final model specification. The Wald test (p < 0.001) provided confirmation that the inclusion of the selected variables enhanced the statistical fit of the two regression models. The LRT ran each of our regression models twice: first without the interaction *sex*rsby* and second with the interaction. The LRT for the GLM regression confirmed significance (p = 0.0994), indicating that including the interaction term improved the final fit of the model. Despite the lack of significance in the LRT for the logistic regression, we included the interaction term in the final model for consistency in data analysis and reporting between the two models. All analyses were conducted using Stata 16.1 (47). Initially, we also intended to analyse women’s health service utilisation and financial protection across the age groups used in this paper. We therefore stratified the bivariate analyses by these age groups, namely 0-14, 15-29, 30-49 and 50+. Results are consistent with those of the main analyses and are included as supplementary files (see supporting information S1 and S2). We also checked whether the outcome variables display different patterns for men and women who had accessed services in public or private facilities. Again, the results (see supporting information S3) are consistent with the main analyses.

### Ethical considerations

Ethical approval for using the original data for the purpose of this study was obtained from the Institutional Review Board of Sigma in New Delhi, India (IRB Number: 10063/ IRB/19-20). Informed written consent was obtained from the study participants before the original data collection was performed. The Ethics Committee of the Medical Faculty of Heidelberg University did not require an ethical approval for this study, as only secondary data were used and no new data collection was carried out. We declare that all methods were performed in accordance with relevant guidelines and regulations.

## 3. Results

Given our analytical focus, we show the complete model results, but report exclusively the results of the main exposure variables *sex* and *enrolment in RSBY*.

### Outcome 1: hospitalisation

Out of the total sample of 36 665 individuals, 51.1% of men and 48.9% of women were enrolled in RSBY. Overall, 1321 (3.6%) individuals were hospitalised, out of which 46.2% were men and 53.8% were women. Out of the hospitalised individuals, 46.8% of men and 53.2% of women were enrolled in RSBY.

When not taking enrolment in RSBY into account, bivariate analyses (Table 3) suggested significant differences between the outcome *hospitalisation* and the exposure variable *sex*. In general, more women were hospitalised than men (women: 4%, men: 3.23%, p < 0.001). However, this difference was not observed for non-MH hospitalisations. Hospitalisation rates were consistently higher among RSBY enrolees, regardless of whether we considered the sample including all services (enrolled: 4.13%, non-enrolled: 3.08%, p < 0.001) or the sample excluding MH services (enrolled: 3.86%, non-enrolled: 2.81%, p < 0.001).

**Table 3:**
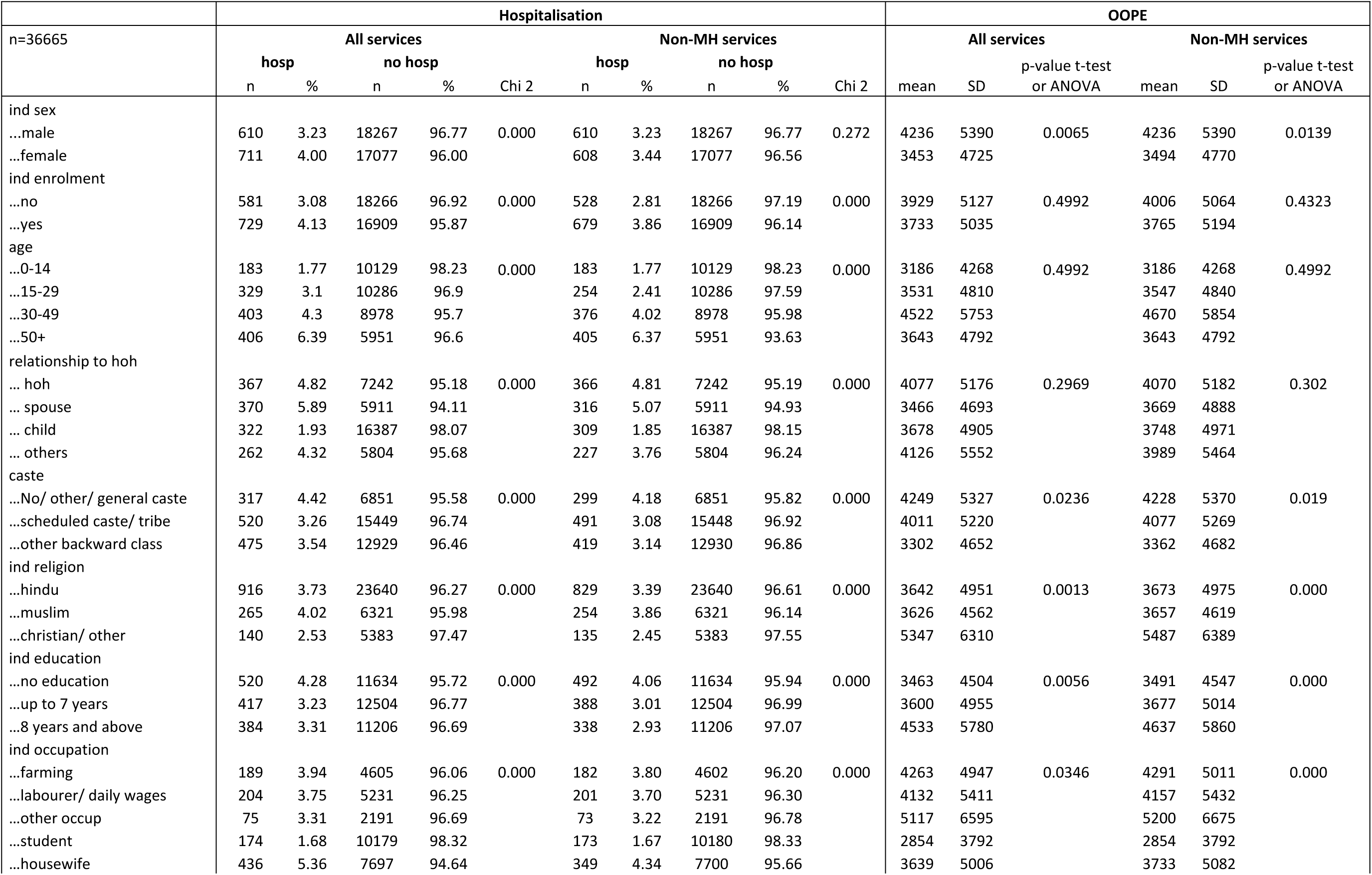
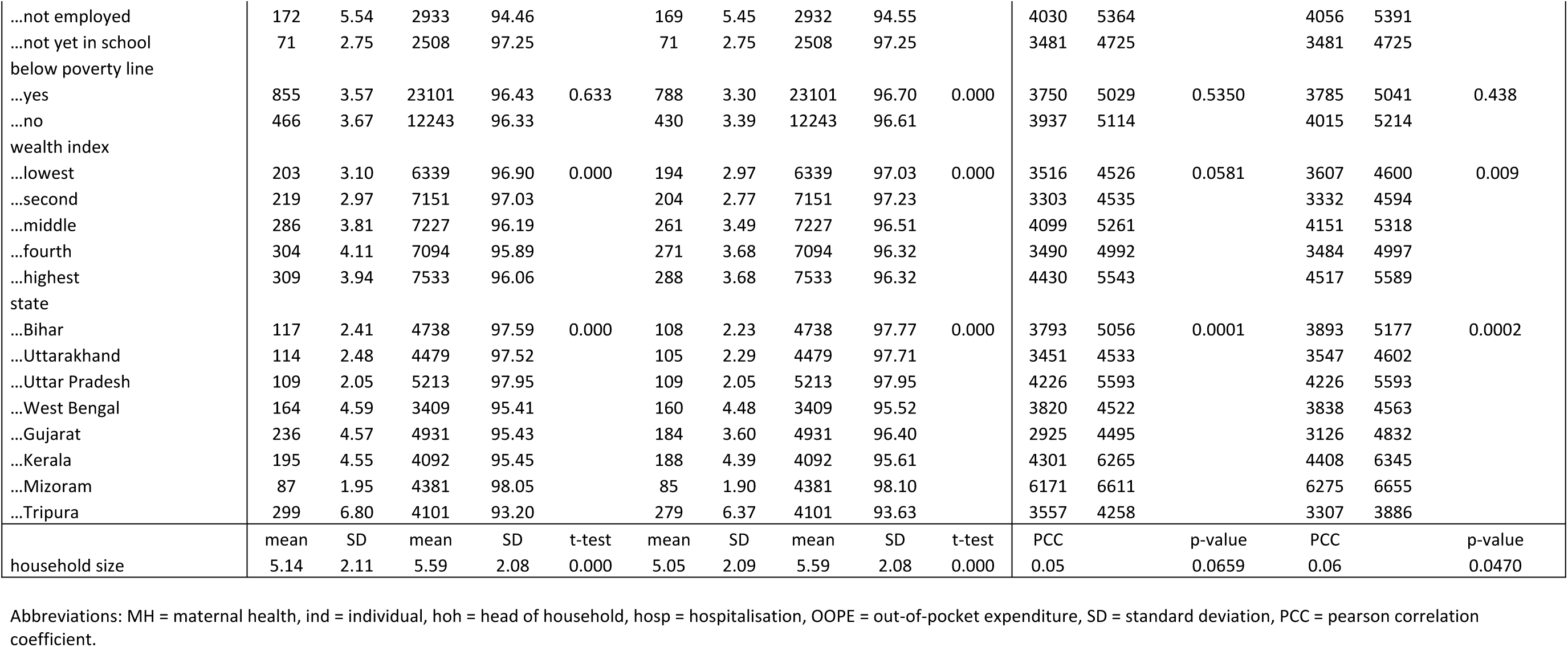
Bivariate analyses for *hospitalisation* and *OOPE*.

Once we differentiated between men and women enrolled and not enrolled in RSBY (Table 4), we observed that more women were hospitalised than men regardless of their enrolment status (enrolled women: 4.5%, enrolled men: 3.78%, p = 0.016; non-enrolled women: 3.48%, non-enrolled men: 2.71%, p = 0.002). Again, this difference was not observed for non-MH hospitalisations.

**Table 4:** Bivariate analysis for hospitalisations of persons enrolled/ not enrolled in RSBY.

| n=36665 | All services |  |  |  |  | Non-MH services |  |  |  |  |
| --- | --- | --- | --- | --- | --- | --- | --- | --- | --- | --- |
|  | hosp |  | no hosp |  | Chi2 | hosp |  | no hosp |  | Chi2 |
|  | n | % | n | % |  | n | % | n | % |  |
| enrolled in RSBY |  |  |  |  |  |  |  |  |  |  |
| ...male | 341 | 3.78 | 8678 | 96.22 | 0.016 | 341 | 3.78 | 8678 | 96.22 | 0.574 |
| ...female | 388 | 4.50 | 8231 | 95.50 |  | 338 | 3.94 | 8231 | 96.06 |  |
| not enrolled in RSBY |  |  |  |  |  |  |  |  |  |  |
| ...male | 265 | 2.71 | 9501 | 97.29 | 0.002 | 265 | 2.71 | 9501 | 97.29 | 0.408 |
| ...female | 316 | 3.48 | 8765 | 96.52 |  | 263 | 2.91 | 8765 | 97.09 |  |

Abbreviations: MH = maternal health, hosp = hospitalisation.

The results from the multivariate logistic regression (Table 5) revealed that there was no statistically significant difference regarding the outcome variable hospitalisation between men and women, even after accounting for enrolment in RSBY. The results were consistent whether one considered the overall hospitalisation sample for all services or the one excluding hospitalisations related to MH services.

**Table 5:**
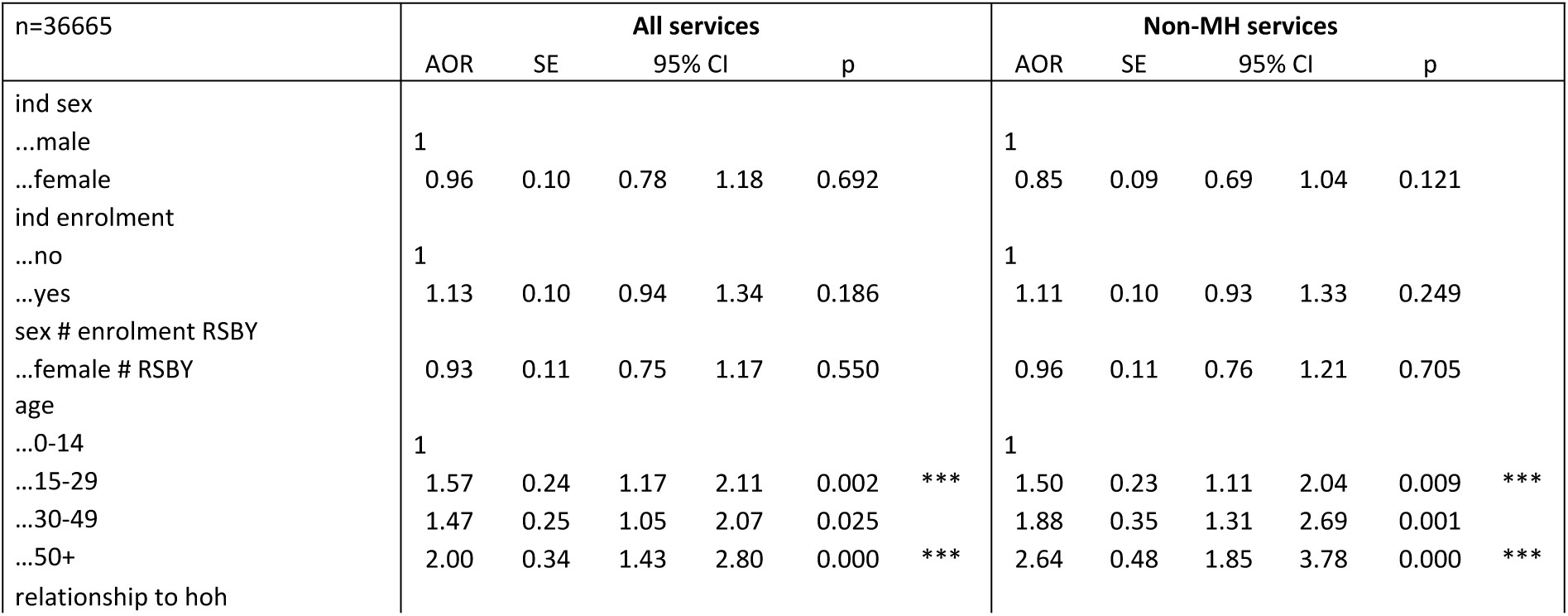
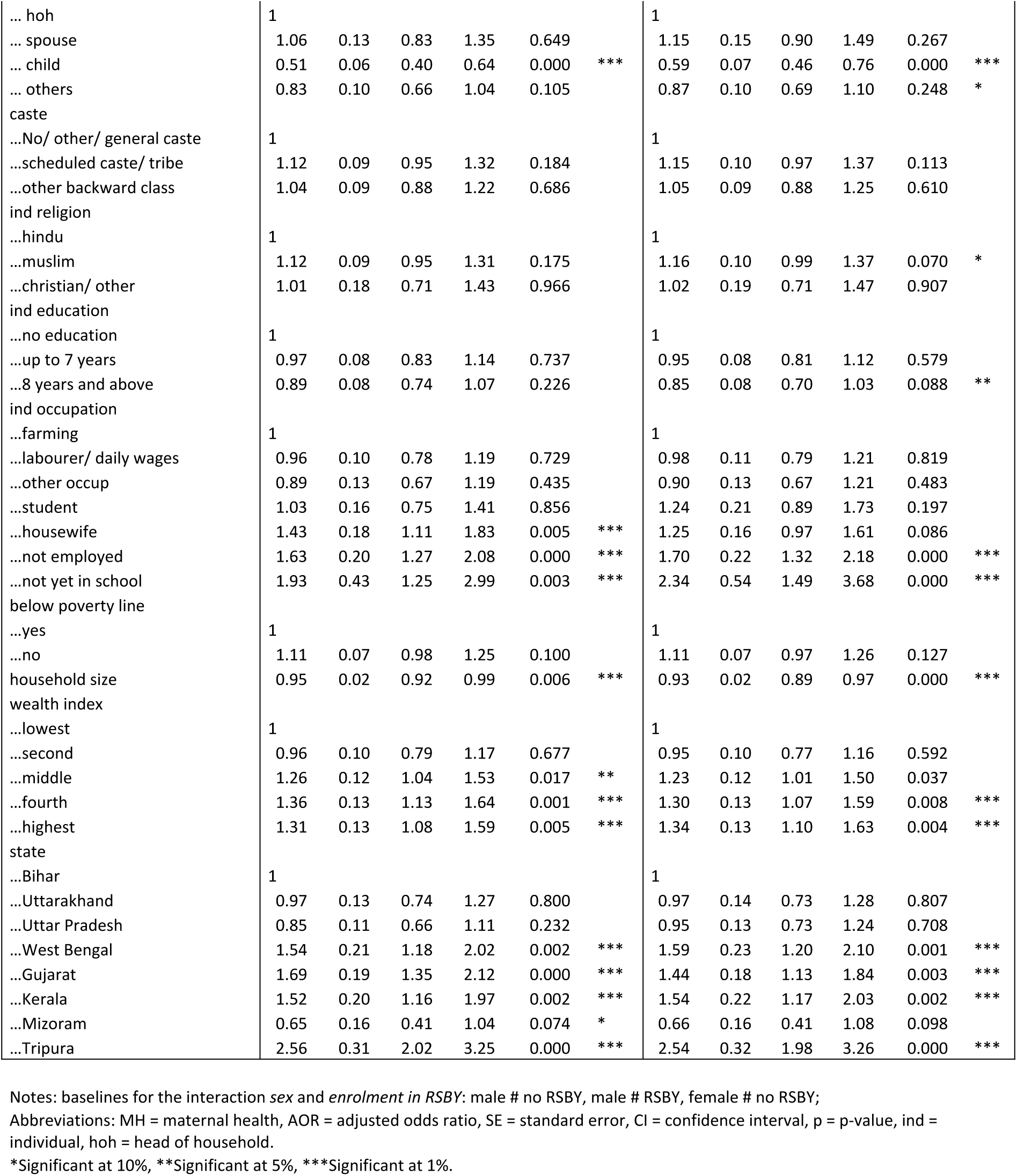
Multivariate logistic regression for *hospitalisation*.

### Outcome 2: OOPE

Out of 1,321 hospitalised, 1,236 (93.57%) individuals experienced OOPE, of which 46.44% were men and 53.56% were women. Among those who incurred OOPE, 663 (53.64%) were enrolled in RSBY, with 46.76% being men and 53.24% being women.

When not considering the RSBY enrolment status of individuals, bivariate analyses (Table 3) suggested significant differences in OOPE based on *sex*: women (mean: 3453 INR, SD: 4725 INR, p = 0.0065) experienced lower OOPE than men (mean: 4236, SD: 5390 INR) for all services as well as for non-MH services (mean: 3494 INR, SD: 4770, p = 0.0139).

Once we differentiated between the enrolment status of men and women, our analysis (Table 6) revealed a similar significant difference in OOPE for individuals in RSBY. Specifically, enrolled women (mean: 3194 INR, SD: 4338 INR, p = 0.0032) experienced lower OOPE than enrolled men (mean: 4346 INR, SD: 5671 INR) for all services and for non-MH services (mean: 3185 INR, SD: 4305 INR, p = 0.0042). This difference disappeared for men and women who were not enrolled in RSBY.

**Table 6:** Bivariate analysis for OOPE of persons enrolled/ not enrolled in RSBY.

| n=1321 | All services |  |  | Non-MH services |  |  |
| --- | --- | --- | --- | --- | --- | --- |
|  | mean | SD | p-value<br>t-test | mean | SD | p-value<br>t-test |
| enrolled in RSBY |  |  |  |  |  |  |
| ...male | 4346 | 5671 | 0.0032 | 4346 | 5671 | 0.0042 |
| ...female | 3194 | 4338 |  | 3185 | 4305 |  |
| not enrolled in RSBY |  |  |  |  |  |  |
| ...male | 4111 | 5087 | 0.4356 | 4111 | 5087 | 0.6407 |
| ...female | 3773 | 5163 |  | 3897 | 5309 |  |

Abbreviations: MH = maternal health, SD = standard deviation.

The results of the GLM model are shown in Table 7. Regarding hospitalisations for all services and without taking RSBY enrolment into account, we observed that OOPE for women (coef = -0.23; p = 0.070) was significantly lower than for men. Looking at the interaction term *sex*rsby*, the results suggest that RSBY did not result in overall reductions in OOPE for the population at large, but it did bring down OOPE for women specifically. While the downward pattern is apparent for hospitalisations for the analysis for all services and for non-MH services, a decrease in OOPE (coef = -0.25; p = 0.087) is significant only for the sample of hospitalisations due to non-MH services.

**Table 7:**
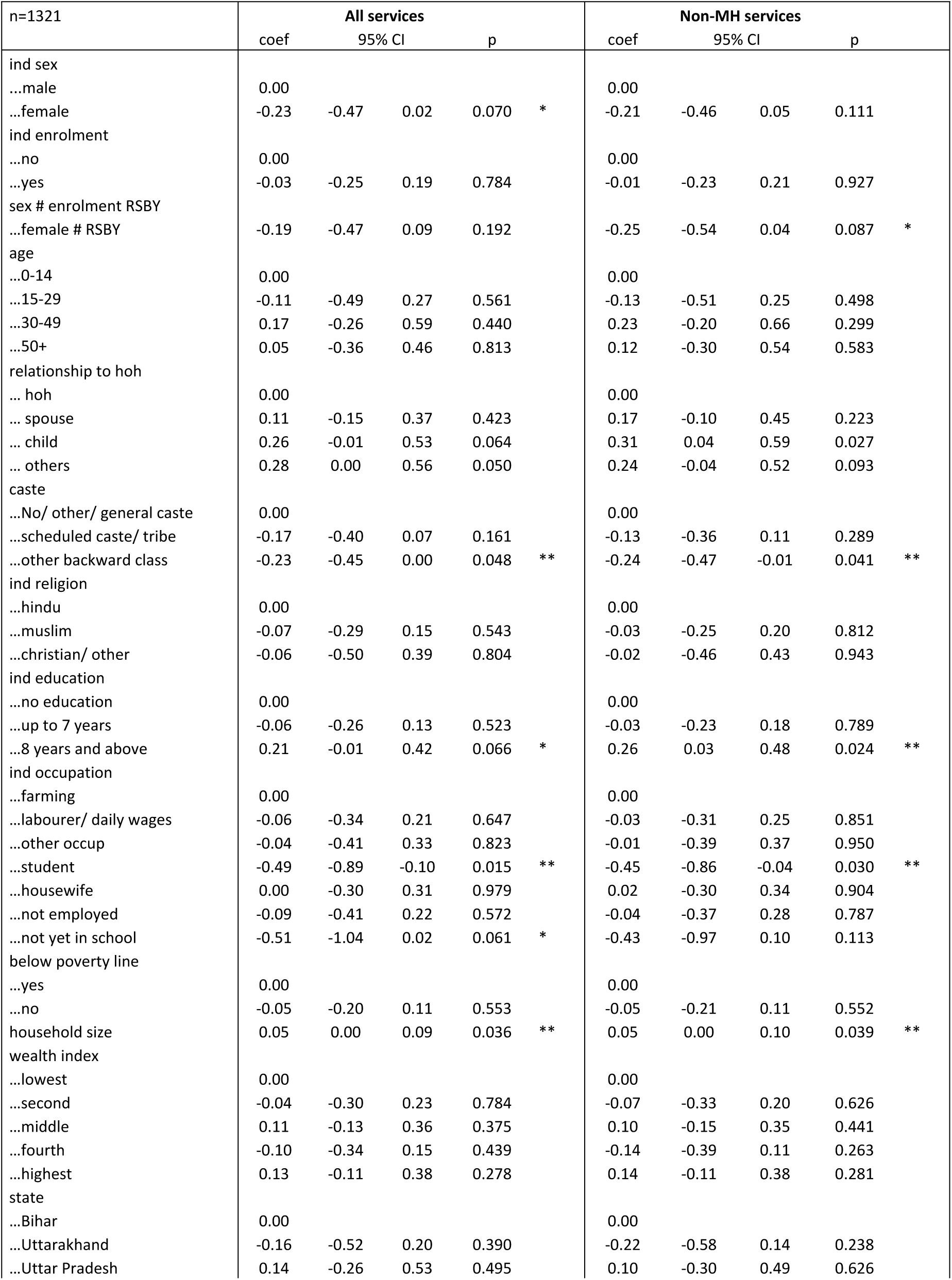
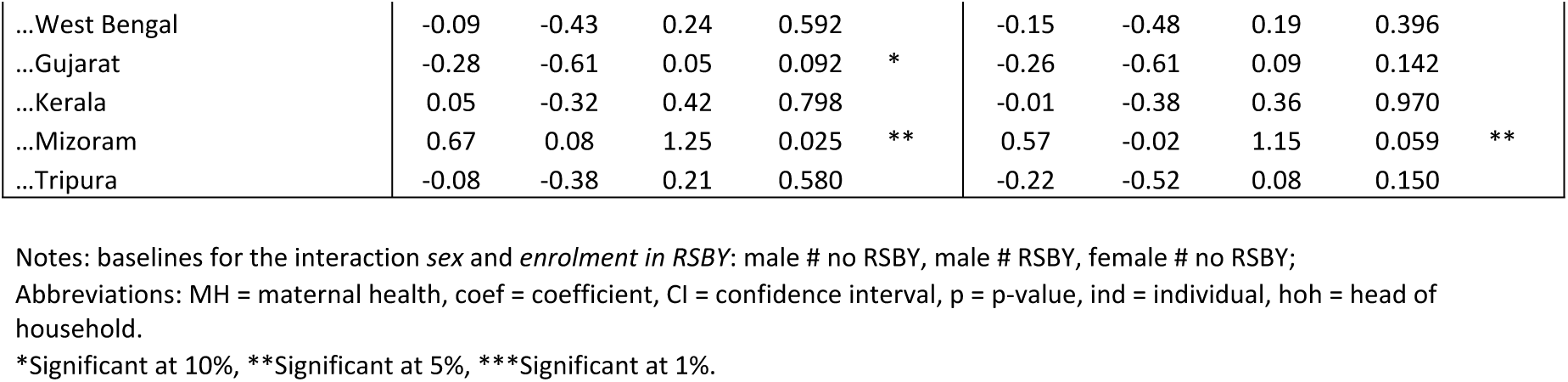
Generalised linear model (GLM) for OOPE.

## 4. Discussion

This study makes an important contribution to the literature by being one of the first to examine differences in utilisation and OOPE between men and women, further distinguishing between MH and non-MH services.

Our key findings reveal, first, that irrespective of enrolment into RSBY, women had a higher use of health care services, but only when MH services were factored into the analysis.

Second, the decision to utilise health care services was not dependent on enrolment in RSBY, for both men and women. And third, RSBY did not decrease OOPE for the population as a whole, but it provided financial protection for women who sought non-MH services. In the following, we discuss these findings in the wider Indian and global socio-economic contexts.

First, we note that – independent of enrolment in RSBY – hospitalisation rates were slightly higher for women than for men, but only if MH services, in our case c-sections and deliveries, were included in the analysis. This result reflects the efforts of the Indian Government to increase institutional deliveries in recent decades (increase from 41% in 2005-06 to 79% in 2015-16) (48). Despite this progress, the use of facility-based care for deliveries in India is still dependent on the socio-economic position and the place of residence, including state and rural-urban locality (48–51).

Our results also revealed that once we excluded MH services from the analysis, the difference in the hospitalisation rate between men and women was lost. This means that, factoring out MH services, hospitalisations under RSBY were largely gender-neutral. A previous study also reported no gender difference in the likelihood of hospitalisations in the general population younger than 60 years, whereas males older than 60 years were more likely to be hospitalised (52). Regarding PM-JAY, general medicine is the most utilised service for men and women, followed by general surgery and oncology for men and by MH services and oncology for women (53). Despite MH services typically driving female enrolment rates and being the second most accessed service by women, women’s overall utilisation of PM-JAY remains lower than for men (48% and 52%) (54). This is evident, for example, in the utilisation of general medicine from 2019 to 2021, where these services accounted for 35.5% among men and 26.2% among women across all services accessed (53). These differences indicate that hospitalisations under PM-JAY still have significant progress to make in achieving gender-equity or neutrality. Nonetheless, our result for RSBY with the difference in hospitalisation rates for women, depending on whether MH services are included or excluded in the analysis, holds important implications for the way how the impact of health policies is evaluated. In order to measure whether a health policy has contributed to improving women’s health, it is essential to expand health utilisation rates beyond maternal health and consider the changing health care needs of both men and women throughout their lifecycles, especially for morbidities where men and women experience similar incidence rates.

Second, our findings reveal that the utilisation of hospitalisation services for men and women was not dependent on enrolment in RSBY. This demonstrates that RSBY has not necessarily contributed to improving access to secondary health care, as enrolment in RSBY did not appear to have led to more hospitalisations. In general, the service uptake of RSBY was low, with a hospitalisation rate ranging between 2% in urban areas and 9% in rural areas, but also higher among women due to the utilisation of MH services (25,27,33,34,55). The novel finding of our study is that the hospitalisation decision under RSBY was largely gender-neutral for enrolled men and women. Regarding enrolment in RSBY, previous studies have already demonstrated a limited or no gender bias and even higher enrolment for female-headed households (27–32)(25,26,56). Evidence from other Indian welfare schemes highlights that despite being enrolled or entitled to benefits, women had difficulties with access because of a lack of decision-making power, restricted mobility and limited access to resources (49,57–59). Maternal empowerment indicators such as the ability to leave the house or economic independence positively correlated with women’s use of MH services under RSBY (36). On the one hand, we assume that the gender-neutrality regarding enrolment and hospitalisation under RSBY might have been the result of pro-women built-in design features of RSBY such as the mandatory enrolment of spouses and the coverage of maternal health services (39,60). On the other hand, the gender-neutral findings suggest that RSBY neither improved nor worsened the conditions for men and women who sought health care services. For a country such as India, where gender disparities in access to health care are well-documented (61–68), this result is an important finding. Early evidence on the implementation of PM-JAY also indicates that having access to the scheme does not lead to increased hospitalisations for either men or women (69). In order to improve access to health services for men and women, learnings from RSBY and other schemes need to be incorporated into both pillars of the Ayushman Bharat Initiative, namely PM-JAY on the one hand and the *Health and Wellness Centres* (HWCs), responsible for delivering primary health care, on the other hand. A gender-sensitive and responsive approach to health needs requires that HWCs and PM-JAY effectively address the health needs of men and women throughout their lifecycles, ensuring that appropriate treatments and health services ranging from primary to secondary and tertiary care are provided at the right time.

Third, more than 93% of men and women in our sample experienced some form of OOPE during hospitalisation. Irrespective of enrolment in RSBY, we observed that OOPE for women was generally lower than for men. We assume that households were inclined to allocate their limited financial resources toward men’s health care rather than women’s or they sought more expensive and higher quality care for men than for women. This is in line with previous studies which demonstrate that households are more likely to rely on OOPE and distressed financing to cover hospitalisation expenses for men rather than women, indicating that India’s gender disparities are also reflected in the financing of health care with large variations by state (65,66,70). When considering enrolment in RSBY in our analyses, we found that RSBY had no effect on OOPE as a result of hospitalisations for the general population. This finding is consistent with studies on RSBY already cited earlier, which underline the limited financial protection RSBY provided to its beneficiaries (27–32). Further analyses revealed that RSBY had an effect and provided financial protection for women who sought non-MH services. This might be explained by the fact that MH services in India are covered by several specific and partially overlapping programs, which might have led women to use RSBY particularly for non-MH services. The financial protection of RSBY for women who sought non-MH services is an important finding and highlights the broader health care needs of women beyond MH. Regarding PM-JAY, research indicates that the scheme did not increase hospitalisation services and also did not reduce OOPE of patients (71). Another study found that PM-JAY has reduced OOPE by 13% for services accessed in private, but not in public facilities, with no differences between men and women (69). These findings highlight the need to further improve the way how PM-JAY is implemented in order to actually ensure financial protection of its beneficiaries. Next to general medicine, MH services are a main driver for women’s overall utilisation of PM-JAY (53), but evidence on the impact of the scheme on OOPE for women’s hospitalisations for MH and non-MH services does not yet exist. Nonetheless, we recommend integrating the finding from RSBY, that a PFHI can financially protect women accessing non-MH services, into a gender-responsive design of services offered under PM-JAY and the HWCs. Firstly, in the set-up of such a design, it is crucial to prevent the misuse of PM-JAY for unnecessary or unintended procedures. For example, in RSBY, hysterectomies have been a common procedure utilised by women, leading to widespread reports of misuse (72). Therefore, for PM-JAY we propose comprehensive counselling on sexual and reproductive health to both women and men as well as eliminating any form of profit incentive for health care facilities to perform unnecessary hysterectomies. Additionally, improved claim reviews and data management are required to assess the necessity of procedures, especially of younger women, as well as variations and distributions of performed procedures within and between states (73). Secondly, when considering our finding alongside evidence from PM-JAY, it emphasises the need for the Indian Government to prioritise policy efforts aimed at promoting equity in the utilisation of services for both men and women, as disparities still exist (74). PM-JAY still lags behind on crucial elements like equity in supply and utilisation, targeting vulnerable population groups, cost coverage and awareness (74,75). And thirdly, our findings can also inform India’s efforts to manage and address the health care needs over the lifecycle. This could include, for example, the set-up of an effective referral system under the Ayushman Bharat Initiative which more closely linking primary, secondary and tertiary care under the HWCs and PM-JAY. Such a referral system would also support the Indian Government’s efforts to fight the growing burden of NCDs by providing comprehensive primary healthcare services through the HWCs (76). The international literature on the role of PFHI in the treatment of non-MH care is limited, including for India. A study using the 2002/03 World Health Survey data found that having all household members enrolled in mandatory or voluntary health insurance improved the likelihood of receiving treatment for chronic care by 38% (77). A study from Brazil showed that individuals with access to health insurance were more likely to use NCD treatments (78).

Taken together, our results suggest that PFHIs can play an important role in ensuring financial protection for women who seek treatment for non-MH services. We recommend additional qualitative research for PM-JAY and other PFHI to understand what causes such effects. This is especially important in the context of discussions on expanding health insurance to the “missing middle”, an estimated number of 300 to 400 million Indians (or 30% of the population) who currently lack any form of either publicly-funded or contributory health insurance (79,80). In addition, women’s access to PFHI in India still very much depends on multiple intersecting determinants like age, education, caste, and wealth (74,81). Findings should feed into reform processes of PFHIs in India and countries worldwide to ensure that these schemes are gender-responsive, equitable and universal in their design and implementation, and ultimately reach all poor and vulnerable. In addition, we recommend similar gender analyses of utilisation and financial protection under PM-JAY, incorporating an intersectional approach that considers factors like age, education, wealth and more.

### Methodological considerations

First, we used secondary data for which districts with high RSBY enrolment rates were purposively selected. This selection might have affected the distribution of enrolment in a non-random way. Second, the available sample for the analysis of hospitalisations was small, which prevents our findings from being generalised to the whole of India or to settings that are different from the study states. Third, we cannot account for the need for hospitalisations, meaning we do not know who in our sample fell sick and who among the sick was hospitalised. Fourth, one could speculate that our analysis is biased as it does not account for the distribution of service use between men and women, hence we cannot attribute observed differences in financial protection to the scheme, but rather to the underlying service use between enrolled men and enrolled women. Our descriptive statistics, however, do not detect major difference in the distribution of service use between enrolled men and enrolled women, hence defeating this argument. Fifth, although hysterectomies have been prevalent in India for years and are a leading reason for women to use health insurance (72), they were not included as a separate hospitalisation category in the household survey used for this study. We recommend research that specifically analyses the use of hysterectomies under PM-JAY. In addition, the survey does not include antenatal or postnatal services as separate categories or a further specification of the categories on gynaecological treatments and on cancer.

Furthermore, although we have tried to account for sex-specific distribution of hospitalisation services, we recognise that due to the self-reported data, 35.3% of services for men and 28.7% of services for women cannot be classified since they were categorised as “Others”. This is a common limitation of self-reported population based data. And sixth, the data were collected in 2014 and RSBY is no longer operational. Nonetheless, the analysis of these data still delivers important practical lessons that can feed into the design and implementation of PFHI in India and other LMICs.

## 5. Conclusion

This study makes an important contribution to the existing literature, as it is one of the first studies to analyse the effect of a PFHI on women’s use of hospitalisation services and related OOPE while differentiating between MH and non-MH services. Our results have important implications: first, PFHI coverage can serve as an important policy tool in promoting NCD and ID treatment in LMICs, particularly for women. Second, if the health outcomes of women are to be improved, it is imperative that studies on PFHIs and other health programs consider utilisation rates that go beyond maternal health. The focus should be the provision of adequate, quality services that cater to the specific needs of both men and women. And third, our results contribute to the evidence-base on gender-responsive health systems and ultimately on achieving UHC. The establishment of a universal health scheme alone is not enough to guarantee universal and equitable access, especially in countries that are characterised by large gender inequalities (3,4,10,13,14). Universal schemes such as PFHIs may unintentionally replicate or even worsen gender inequalities in health care access, unless the schemes explicitly account for tackling gender inequalities that exist at the societal level through a gender-responsive design and implementation (3,10,13,14,82,83). The results from this paper demonstrate that such a design needs to be integrated into PFHIs in India if access to health care is to be improved, especially for women and girls.

## Data Availability

The original data are available with the Deutsche Gesellschaft für Internationale Zusammenarbeit (GIZ) GmbH in New Delhi, India. GIZ carried out the original data collection on behalf of the Indian Government (Ministry of Labour and Employment). The data are not publicly available. The authors obtained written permission from GIZ to use the data for the purpose of this study and GIZ shared the data with the authors in fully anonymised format. Data may be shared on request to the corresponding author with permission of GIZ.

## Acknowledgments

We would like to thank GIZ India for providing the original data, Prognosis Management & Research Consultants Pvt. Ltd. and NR Management Consultants India Pvt. Ltd. for their efforts in collecting the data as well as the many men and women in India who participated in the original study and contributed their valuable time and insights.

